# Previous Infection Combined with Vaccination Produces Neutralizing Antibodies With Potency Against SARS-CoV-2 Variants

**DOI:** 10.1101/2021.08.27.21262744

**Authors:** F. Javier Ibarrondo, Christian Hofmann, Ayub Ali, Paul Ayoub, Donald B. Kohn, Otto O. Yang

**Affiliations:** Division of Infectious Diseases, Department of Medicine, David Geffen School of Medicine at University of California Los Angeles, Los Angeles, CA 90095; Department of Microbiology, Immunology, and Molecular Genetics, David Geffen School of Medicine at University of California Los Angeles, Los Angeles, CA 90095; Division of Pediatric Hematology Oncology, David Geffen School of Medicine at University of California Los Angeles, Los Angeles, CA 90095

**Keywords:** SARS-CoV-2, COVID-19, vaccines, humoral immunity, spike variants, vaccine efficacy

## Abstract

SARS-CoV-2 continues to evolve in humans. Spike protein mutations increase transmission and potentially evade antibodies raised against the original sequence used in current vaccines. Our evaluation of serum neutralizing activity in both persons soon after SARS-CoV-2 infection (in April 2020 or earlier) or vaccination without prior infection confirmed that common spike mutations can reduce antibody antiviral activity. However, when the persons with prior infection were subsequently vaccinated, their antibodies attained an apparent biologic ceiling of neutralizing potency against all tested variants, equivalent to the original spike sequence. These findings indicate that additional antigenic exposure further improves antibody efficacy against variants.

**LAY SUMMARY:** As SARS-CoV-2 evolves to become better suited for circulating in humans, mutations have occurred in the spike protein it uses for attaching to cells it infects. Protective antibodies from prior infection or vaccination target the spike protein to interfere with its function. These mutations can reduce the efficacy of antibodies generated against the original spike sequence, raising concerns for re-infections and vaccine failures, because current vaccines contain the original sequence. In this study we tested antibodies from people infected early in the pandemic (before spike variants started circulating) or people who were vaccinated without prior infection. We confirmed that some mutations reduce the ability of antibodies to neutralize the spike protein, whether the antibodies were from past infection or vaccination. Upon retesting the previously infected persons after vaccination, their antibodies gained the same ability to neutralize mutated spike as the original spike, suggesting that the combination of infection and vaccination drove the production of enhanced antibodies to reach a maximal level of potency. Whether this can be accomplished by vaccination alone remains to be determined, but the results suggest that booster vaccinations may help improve efficacy against spike variants.

## BACKGROUND

Evolution of the SARS-CoV-2 spike protein results from selection for random mutations that yield fitness benefit that promotes person-to-person dissemination of the virus. Mutations in the receptor-binding domain (RBD), which is involved in viral entry and the key target of neutralizing antibodies, may also facilitate viral resistance to antibodies against the original spike sequence either from prior infection or the current vaccines. Spike variants such as B.1.351/*Beta* [1] and P.1/*Gamma* [2] have become predominant over the original sequence of the virus in some geographic locations, and a growing body of evidence suggests that some mutants can contribute to reinfections [3] and vaccine failures [4, 5].

Here we examine the impact of several commonly observed spike mutations on the serum neutralizing activity of persons who had COVID-19 in April 2020 or earlier, of vaccinated persons who had no prior history of infection, and of the COVID-19 recovered persons after subsequent vaccination. This examination directly compares serum neutralization of seven commonly observed spike variant combinations of five RBD mutations by antibodies elicited by vaccination alone, SARS-CoV-2 infection alone, and combined infection and vaccination, to evaluate whether repeated antigenic exposures would increase efficacy of humoral immunity to cope with spike mutations.

## METHODS

### Participants

Volunteers provided informed consent under an institutional review board-approved protocol. Persons with mild SARS-CoV-2 infection were evaluated shortly after recovery from illness, and these persons as well as healthy control individuals without prior SARS-CoV-2 infection were evaluated soon after vaccination for COVID-19. The persons with prior infection all had symptom onset no later than April 2020 in the USA, when the SARS-CoV-2 variants were not yet reported to circulate, and all had mild infection that did not require hospitalization.

### Serum ELISA for anti-RBD antibodies

Anti-RBD antibodies were evaluated with a quantitative ELISA as recently described in detail [6, 7]. In brief, we utilized a modification of a previously reported ELISA [8-10] against recombinant receptor binding domain (RBD) protein bound to a 96-well microtiter plate, using secondary goat anti-human IgG-, IgM-, or IgA-horseradish peroxidase-conjugated detector antibodies. For quantitation, each plate contained serial dilutions of the CR3022 monoclonal antibody in IgG, IgM, or IgA formats, and the amount of binding activity in serum was expressed as an equivalent amount of the control antibody.

### Serum spike-neutralizing assay and definition of neutralizing potency

Spike neutralization activity was assessed with a spike-pseudotyped lentiviral vector assay as recently described in detail [7]. Briefly, an HIV-1-based lentiviral vector delivering a luciferase reporter gene was pseudotyped with the indicated spike variant. The assay was performed by adding serial dilutions of serum during infection of ACE2-expressing HEK 293T cells and assessing luciferase activity after 48 hours. The Hill equation was utilized to estimate the dilution factor yielding 80% inhibition (DF_80_). The “neutralizing potency” (neutralizing activity adjusted for antibody concentration) was defined as the ratio of neutralization activity (DF_80_) to anti-RBD antibody concentration (log_10_ sum of IgG, IgM, and IgA anti-RBD antibodies in ng/ml), adjusting neutralizing activity for antibody quantity.

## RESULTS

### Mutations in RBD variably reduce the spike neutralizing activity of antibodies elicited by vaccination

Serum spike-neutralizing activity was assessed using a spike-pseudotyped neutralization assay and compared to the amount of anti-RBD antibodies to yield a measurement of antibody neutralizing potency. In 15 persons without a history of prior infection who were recently vaccinated (seven with BNT162b2 and eight with mRNA-1273), potency against spike variants was variably less than against the original sequence contained in the vaccine (Figure 1A). The mutants with most reduced potency were K417N/E486K/N501Y and K417T/E486K/N501Y found in B.1.351/*Beta* [1] and P.1/*Gamma* [2] respectively. In contrast to a prior report [5], K501Y found in B.1.1.7/*alpha* exhibited no appreciable resistance to neutralization. There were no clear differences between participants receiving different vaccines, although there were too few subjects for meaningful comparisons. Overall, these results demonstrated that spike mutation can affect the sensitivity to neutralization by antibodies elicited by vaccination with the original spike sequence.

**Figure 1.**
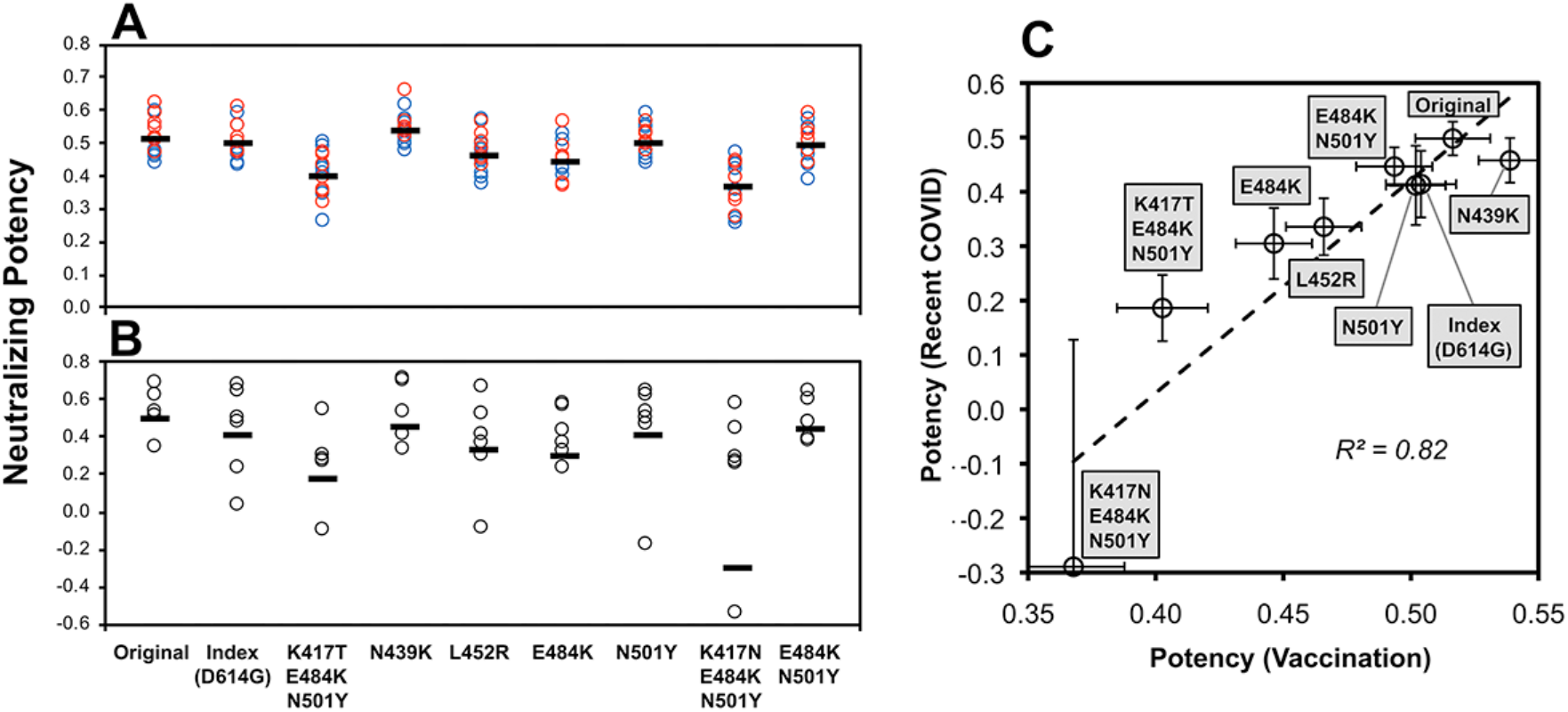
SARS-CoV-2 spike variant neutralizing potency of antibodies generated by vaccination versus natural infection. Potency of serum antibodies against RBD was estimated as the ratio of serum neutralizing activity (log_10_ dilution factor yielding 80% neutralization of spike-mediated luciferase-expressing lentiviral entry of ACE2-expressing 293T cells) to the concentration of anti-RBD antibodies (log_10_ ng of summed IgG, IgM, and IgA determined by quantitative ELISA using CR3022-monoclonal antibody-based standards) for each of the indicated variants. Potencies were determined against a panel of spike variants containing the indicated mutations; “Original” indicates the unmodified Wuhan-Hu-1 sequence (GenBank #MN908947.3). “Index” is the original sequence with the D614G mutation and the seven other variants are modifications of the index sequence (containing D614G). A. Serum antibody neutralizing potencies are shown for 15 persons without prior SARS-CoV-2 infection after recent vaccination 7 to 26 days after completing vaccination, mean 17 days. Seven received BNT162b2 (red) and eight received mRNA-1273 (blue) vaccines. B. Serum antibody neutralizing potencies are shown for 10 persons with recent mild SARS-CoV-2 infection (18 to 45 days after onset of symptoms, mean 31 days). These persons were all infected in the United States prior to May 2020, before substantial prevalence of spike variants appeared. C. The means and standard errors for potencies against each variant for each group (vaccinees versus recent COVID-19) are plotted against each other.

### Mutations in RBD also variably reduce the spike neutralizing activity of antibodies elicited by natural infection with a similar pattern

Serum antibody spike-neutralizing potency was also assessed for 10 persons with recent mild SARS-CoV-2 infection (mean 31 days after onset of symptoms, range 18 to 45 days) who were infected in the United States prior to May 2020, before reports of circulating variants. Again, potency was variably affected by different mutations, and the mutants with most reduced neutralizing potency were K417N/E486K/N501Y and K417T/E486K/N501Y (Figure 1B). The neutralizing potencies of these infected persons correlated to those of the previously uninfected vaccinees, indicating similar susceptibilities to spike mutation of antibodies raised by either natural infection or vaccination alone.

### Vaccination of previously SARS-CoV-2-infected persons increases neutralizing potency against variants to an apparent biologic ceiling

When the prior-infected individuals were vaccinated after approximately a year (mean 353 days after symptom onset, range 280 to 394 days), serum neutralizing potency against the original sequence spike was relatively unchanged, but activity against all variants equalized to a level similar to the original sequence (Figure 2A). Neutralizing potency change correlated tightly to the baseline early after infection across all individuals and variants (Figure 2B), suggesting a consistent biologic ceiling across all variants. Again, there were no clear differences between persons receiving different vaccines. Overall, these findings indicated that the combination of natural infection and vaccination drove antibody potency to an apparent maximum for all tested variants.

**Figure 2.**
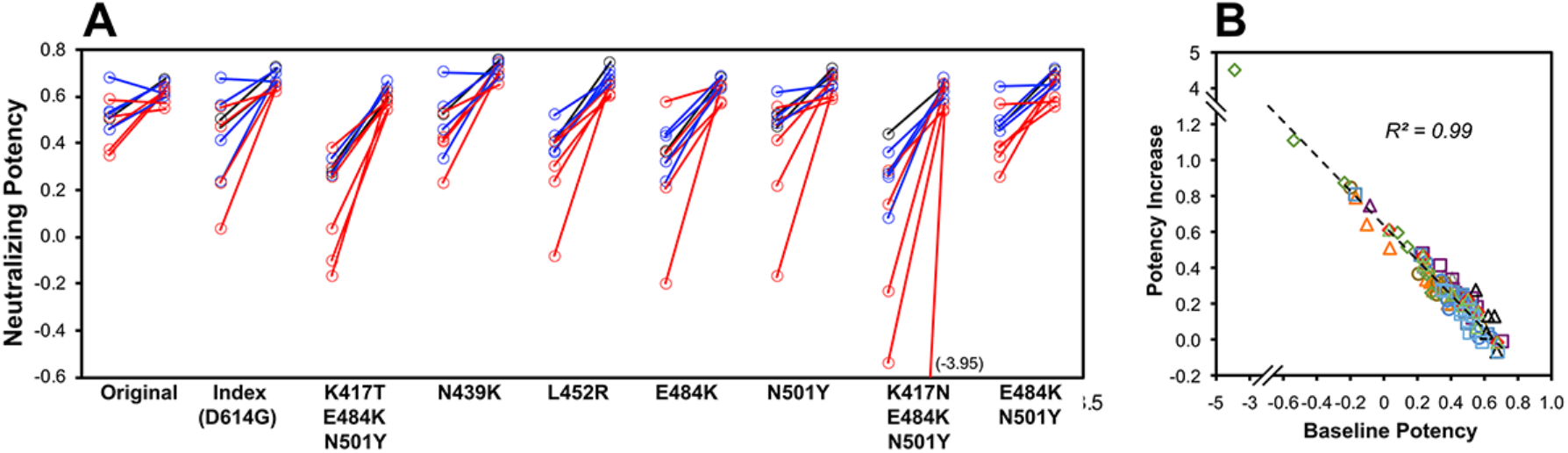
Changes in SARS-CoV-2 spike variant neutralizing potency of antibodies after vaccination of persons with prior SARS-CoV-2 infection. For the 10 persons with prior COVID-19 described in Figure 1, vaccination was initiated a mean of 353 days after symptom onset (range 280 to 394 days) and serum neutralizing potency against each variant was determined after vaccination. A. Potency against each variant is plotted consecutively for recent infection (18 to 45 days after onset of symptoms, mean 31 days) and post-vaccination. Five persons received BNT162b2 (red), four persons received mRNA-1273 (blue), and one person received Ad26.COV2.S (black). B. The changes in potencies against each variant are plotted against the initial potencies against each corresponding variant. Each different symbol depicts one variant.

## DISCUSSION

Our results confirm that vaccine- or infection-elicited antibodies raised against the original sequence can lose activity against recently circulating spike variants, substantiating evidence of reduced vaccine protection against variants, such as Vaxzevria and BNT162b2 against B.1.351/*Beta* [4, 5, 11]. Furthermore, our findings indicate that both mild natural infection and vaccination presenting the original spike sequence elicit antibodies with similar patterns of susceptibility to variants.

After subsequent vaccination, however, antibodies in previously infected persons gain spike neutralizing potency against all tested variants equal to the original sequence, although activity against the original sequence itself is relatively unchanged. This suggests that 1) potency against the original sequence was already maximal, and 2) the additional antigenic exposure from vaccination further increased antibody potency against suboptimally recognized variants to reach the same maximal potency.

This gained ability is compatible with further maturation of humoral immunity after SARS-CoV-2 infection and vaccination, consistent with reports of ongoing affinity maturation after resolved infection [12-14]. It is unclear whether our observation relates to time elapsed after infection as proposed by that report, additional maturation due to vaccination, or both. In contrast to studies showing ongoing affinity maturation, a recent report suggests that potency declines after resolution of infection [7], suggesting that vaccination has a key role in this phenomenon. Finally, the apparent ceiling for antiviral potency against all tested variants is consistent with attaining a maximum threshold of binding affinity [15, 16] for the original sequence that eventually also applies to the tested variants.

Overall, our findings raise the possibility that resistance of SARS-CoV-2 spike variants to antibodies can be overcome by driving further maturation through continued antigenic exposure by vaccination, even if the vaccine does not deliver variant sequences. Whether this can also be accomplished in SARS-CoV-2-naïve persons through vaccination alone, such as delivering booster doses beyond the original vaccination regimen of two doses, remains to be determined.

## Data Availability

Provided upon request

## ACKNOWLEDGEMENTS

We are grateful for the generous participation of the volunteers who donated their blood to these studies.

